# Neuroimaging Insights into Brain Mechanisms of Early-onset Restrictive Eating Disorders

**DOI:** 10.1101/2024.11.12.24317128

**Authors:** Clara A. Moreau, Anael Ayrolles, Christopher R. K. Ching, Robin Bonicel, Alexandre Mathieu, Coline Stordeur, Pierre Bergeret, Nicolas Traut, Lydie Tran, David Germanaud, Marianne Alison, Monique Elmaleh-Bergès, Stefan Ehrlich, Paul M. Thompson, Thomas Bourgeron, Richard Delorme

## Abstract

**Background:** Early-onset restrictive eating disorders (rEO-ED) encompass a heterogeneous group of conditions, including early-onset anorexia nervosa (EO-AN) and avoidant restrictive food intake disorders (ARFID). Almost nothing is known about the consequences of rEO-ED on brain development.

**Methods:** We performed the largest comparison of MRI-derived brain features in children and early adolescents (<13 years) with EO-AN (n=124), ARFID (n=50), and typically developing individuals (TD, n=112).

**Results:** Despite similar body mass index (BMI) distributions, EO-AN and ARFID showed divergent structural patterns, suggesting independent brain mechanisms. Half the regional brain measures were correlated with BMI in EO-AN and none in ARFID, indicating a partial mediation of EO-AN signal by BMI. EO-AN was associated with a widespread pattern of thinner cortex, while underweight ARFID patients exhibited smaller surface area and subcortical volumes than TD.

**Conclusion:** Future studies will be required to partition the contribution of low BMI *vs.* ED mechanisms in neurodevelopmental disorders.

## Introduction

Early-onset restrictive eating disorders (rEO-ED) in children encompass a heterogeneous group of conditions, including early-onset anorexia nervosa (EO-AN) and avoidant restrictive food intake disorder (ARFID). Both disorders are characterized by severe food restriction and significant weight loss but differ in the primary drivers of restrictive behavior ^1,2^. Pediatrics national surveillance studies have reported an increasing prevalence of rEO-ED, ranging from 1.4 to 3/100 000, with an incidence doubling over the past decade ^3–6^. The COVID-19 pandemic also contributed to a strong rise in ED incidence, with a concomitant increase in healthcare costs ^7,8^.

Early-onset form of Anorexia Nervosa (symptom onset <13 years old, EO-AN) has been associated with greater detrimental impacts on bone development, statural growth, and puberty, as well as faster weight loss compared to patients with a typical onset (TO-AN) ^9^. Patients with EO-AN show distinct epidemiological and clinical features compared to TO-AN, including a more balanced sex ratio (3 boys for 7 girls vs. 1 for 9 in TO-AN), a higher frequency of the restrictive subtype (rare binge-purge symptoms), a lower incidence rate, and more rapid weight loss ^9^. Despite these differences with TO-AN, very little is known about the mechanisms of EO-AN and its impact on brain development. The largest study to date of TO-AN was conducted by the ENIGMA (Enhancing Neuro Imaging Through Meta-Analysis) Eating Disorders working group, which included 685 AN and 963 typically developing individuals (TD) (mean age at scan = 21 [15-27] years with a mean illness duration of 5 [1-13] years) ^10^, and revealed a widespread pattern of lower cortical thickness (CT) and smaller thalamic volumes in TO-AN, relative to TD. Brain alterations were less pronounced in weight-restored individuals, suggesting a mediating effect of body mass index (BMI) on brain structure in TO-AN ^10^.

The extent to which brain alterations in AN are driven by poor nutrition and low BMI or represent distinct AN-related pathophysiological processes remains largely unknown. One way to tackle this question would be to investigate the effect of low BMI in individuals with restrictive dietary behaviors but without AN. ARFID is a rEO-ED condition characterized by a limitation in the amount or type of food consumed that is not motivated by body image concerns or fear of gaining weight ^11^. Three clinical subgroups have been described: lack of food interest, fear of aversive consequences, and sensory sensitivity impairment. Prevalence estimates are elevated and vary across studies up to 15% in general population, including both selective and restrictive forms. The prevalence of restrictive forms remains unclear ^12^. To date, no studies have evaluated the impact of ARFID on brain structure. As a significant proportion of ARFID patients are severely underweight (BMI<3rd percentile), comparing ARFID patients with EO-AN could provide a way of disentangling brain alterations driven primarily by low BMI and those resulting from EO-AN pathophysiology.

Clinical observations (overlapping symptoms and comorbidities) and genetic analyses (genetic correlations of risk variants) have highlighted similarities between AN and other serious mental illnesses ^13,14^. Almost one-fifth of AN patients meet criteria for obsessive-compulsive disorder (OCD)^14,15^. Overlapping symptoms and comorbidities with autism spectrum disorder (ASD)^16,17^, major depression disorder (MDD), and anxiety disorders are also commonly observed in AN ^13,15^. At the genetic risk level, variants associated with AN derived from genome-wide association studies (GWAS) show significant correlations with variants associated with OCD (r_G_=0.45) and MDD (r_G_=0.27) ^14^. Similarly, clinical observations in ARFID have revealed an association with anxiety disorders and neurodevelopmental disorders (NDD), including ASD and attention deficit-hyperactivity disorder (ADHD) ^1,12,18^. Recent GWAS have revealed a high heritability of ARFID^19,20^, but nothing is known so far about similarities with other psychiatric conditions at the genetic level. The extent to which rEO-ED displays overlapping or distinct brain signatures compared to other clinically and genetically associated mental illnesses is currently unknown.

We investigated three main aims:

(1) What structural brain differences distinguish EO-AN from TD, and to what extent does the age of onset of AN influence brain development? ***Hypothesis:*** Individuals with EO-AN will show a widespread pattern of thinner cortex compared to TD. EO-AN will exhibit greater effect sizes than patients with TO-AN ^10^.
(2) Do structural brain profiles differ between EO-AN and ARFID in patients matched for BMI? How does BMI relate to the degree of brain variation in EO-AN? And what structural brain traits differentiate ARFID from TD? ***Hypothesis:*** Underweight patients with ARFID would have a unique pattern of brain abnormalities, partially similar to AN-associated brain abnormalities. This shared brain pattern may reflect a general consequence of low BMI on brain morphometry.
(3) To what extent do brain alterations in rEO-ED overlap with other neurodevelopmental disorders? ***Hypothesis:*** Brain abnormalities in rEO-ED will significantly overlap with brain profiles previously associated with clinically similar conditions.

To this end, we collected and analyzed brain imaging data in children and young adolescents with an early-onset form of AN (<13 y) (n=124), ARFID (n=50), as well as TD (n=112).

## Results

### Widespread pattern of thinner cortex and smaller subcortical volumes in EO-AN

#### Global effects

A widespread global pattern of thinner cortex (Cohen’s *d*=-0.80, p_adj_=2.7e-08), lower total GV (Cohen’s *d*=-0.56 p_adj_=5.4e-0.5), and cerebrospinal fluid (CSF) volume increase (Cohen’s *d*=0.75, p_adj_=9.9e-08) were observed in patients with EO-AN compared to TD (**Figure 1A**, Supplementary Table). We found no significant effect on the total surface area (SA) (Cohen’s *d*=-0.18, ns) or the intracranial volume (ICV) (Cohen’s *d*=0.13, ns) in patients with EO-AN compared to TD.

**Figure 1.**
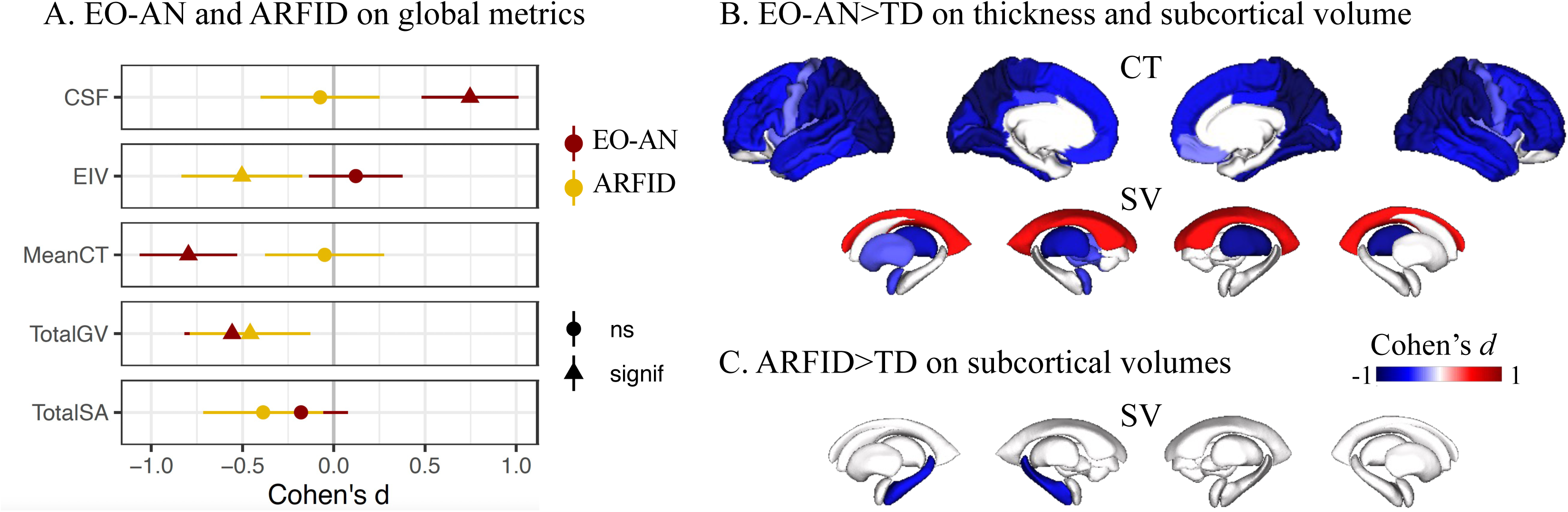
Characterization of rEO-EDs’ effect on brain structure. *A.* Effect of EO-AN (dark red dots) and ARFID (yellow dots) on five global brain metrics (ICV: intracranial volume, Total SA: total surface area, Mean CT: mean cortical thickness, CSF: cerebrospinal fluid, Total GV: total gray matter volume). Triangles represent significant effect sizes. B-C. Brain maps (FDR thresholded) showing Cohen’s d values for each of the 68 cortical regions (thickness) and the 16 subcortical regions and ventricles (volumes) for EO-AN versus typically developing individuals (TD) (B) and ARFID versus TD. CT: cortical thickness, SV: subcortical volume.

#### Regional effects

A widespread pattern of lower cortical thickness was identified, with a stronger contribution observed in the parietal and occipital lobes bilaterally. Fifty-six regions showed significant alterations (**Figure 1B**, Supplementary Table). The regions with the largest effect sizes indicating thinner cortex included the left superior parietal cortex (Cohen’s d= −1.12, p_adj_=1.3e-13) and the precuneus (Cohen’s d=-1.01, p_adj_=1.2e-11) along with the lateral occipital (Cohen’s d=-0.98, p_adj_=2.5e-11). Results adjusted for mean thickness are reported in the Supplementary Results (Figure S3A). No significant regional alterations were found for SA (Supp Table), so we did not include SA metrics in subsequent analyses. At the subcortical level, analyses comparing EO-AN and TD demonstrated smaller bilateral thalamic volumes (Cohen’s *d*=-0.83 and −0.70 for the right and left side, respectively, p_adj_=2.4e-08 and p_adj_=1.7e-06), left amygdala (Cohen’s *d*=-0.40, p_adj_=9.4e-03), pallidum (Cohen’s *d*=-0.37, p_adj_=0.02), and putamen (Cohen’s *d*=-0.34, p_adj_=0.02). We also observed larger bilateral ventricle volumes (Cohen’s *d*=0.50, p_adj_=7.6e-04) (**Figure 1B**, Supplementary Table).

We found an even stronger effect on CT (thinner) and subcortical volumes but still no effect on SA in the acutely ill subgroup of EO-AN patients (EO-acAN, BMI<3rd percentile), compared to TD (**Figure 2A**, Supplementary Figure S5).

**Figure 2.**
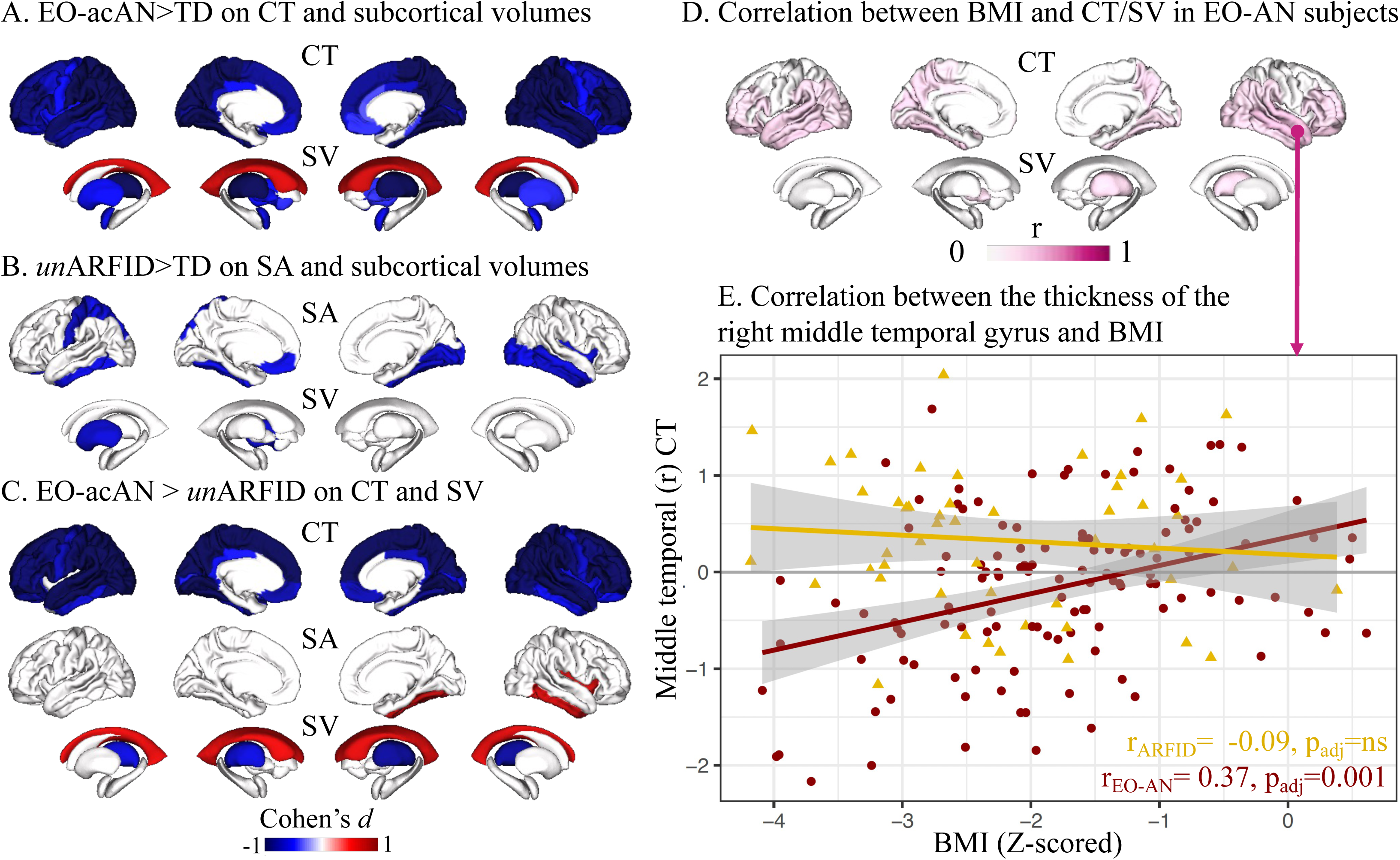
Partition of BMI vs. rEO-ED effects on brain features. *A.* Effects on cortical thickness (CT) and subcortical volumes (SV) of EO-acAN (acutely-ill subgroup, BMI <3rd percentile at scan) versus TD. *B.* Effects on surface area (SA) and SV of unARFID (underweight subgroup, BMI <3rd percentile at scan) versus TD. *C.* Effects of the EO-acAN versus unARFID on CT, SA and SV. *D.* Brain maps summarizing the 84 correlations between BMI and CT (68 cortical regions) as well as SV (14 subcortical regions + 2 lateral ventricles) in EO-AN patients. *E.* Correlations between residuals (removing the effect of age, sex, and machine) of the thickness of the right middle temporal gyrus and Z-scored BMI in individuals with EO-AN (dark red dots) and ARFID (yellow dots). We selected the right middle temporal gyrus as it showed the highest correlation with BMI in patients with EO-AN.

As most prior neuroimaging studies of AN focused on female populations, we conducted similar analyses excluding male individuals and observed consistent results with those obtained in the whole study population (*r*=0.97). We also conducted similar analyses excluding 23 subjects with psychotropic medications (see Supplementary Method) and found no significant difference (*r*=0.99).

### No major differences in brain patterns associated with early vs. typical onset forms of AN

We found a correlation between EO- and TO-AN ^10^ brain profiles at r=0.76 (CI: 0.64-0.85, p=3.5e-14) for the thickness of the 68 cortical regions and r=0.82 (CI: 0.55-0.93, p=9.6e-05) for the volume of the 16 subcortical regions.

The main differences between EO-AN and TO-AN were observed in effect sizes for specific brain regions. For example, the transverse temporal gyrus showed a larger effect in EO-AN>TD (−0.85) compared to TO-AN>TD (−0.18), while the superior frontal gyrus had a greater effect in TO-AN>TD (−0.85) than EO-AN>TD (−0.58) - see Supplementary Figure 4.

Comparison of the distributions of regional effect sizes showed a larger effect on CT in EO-AN>TD (absolute mean top-decile=0.99) compared to TO-AN>TD (absolute mean top-decile=0.89), and a larger effect on subcortical volumes in EO-AN>TD (absolute mean top-decile=0.83) compared to TO-AN>TD (absolute mean top-decile=0.66).

Overall, these differences between EO-AN and TO-AN remain small and suggest a high degree of replication across independent cohorts - as well as signal stability on CT and subcortical volume metrics in AN, independent of the age of onset.

### Distinct structural brain patterns associated with ARFID and EO-AN diagnoses

ARFID was associated with smaller ICV (Cohen’s *d*=-0.50, p_adj_=0.02), total GV (Cohen’s *d*=-0.46, p_adj_=0.03), and left hippocampal subcortical volume (Cohen’s *d*=-0.58, p_adj_=0.02) compared to TD (**Figure 1AC**, Supplementary Table). No effect on CT or SA at the global or regional levels was observed in ARFID>TD. When investigating specifically the underweight group of patients with ARFID (*un*ARFID, BMI<3rd percentile), we observed a large effect on ICV (Cohen’s *d*=-0.70, p_adj_=0.003), total GV (Cohen’s *d*=-0.69, p_adj_=0.003) and a reduction of SA (Cohen’s *d*=-0.59, p_adj_=0.01) (see Supplementary Figure 5). At the regional level, we observed lower SA in 12 cortical regions (**Figure 2.B**). The top impacted regions were the left postcentral area (Cohen’s *d*=-0.72, p_adj_=0.02) and the right fusiform gyrus (Cohen’s *d*=-0.70, p_adj_=0.02). Finally, we observed smaller putamen volume (Cohen’s *d*=-0.67, p_adj_=0.03) in *un*ARFID>TD (**Figure 2.B**).

To disentangle the effects of the low BMI vs. the ED mechanisms, we compared patients with EO-acAN vs. patients with *un*ARFID (both groups with a BMI percentile<3 at scan) and observed a similar profile to the one observed when comparing EO-acAN vs. TD for thickness and volumes (r=0.86, p<2.2e-16 and r=0.83, p=5.3e-05 respectively, see **Figure 2C**), but not for SA (r=0.33, p=0.005). These analyses demonstrate independent brain structural profiles associated with ARFID and EO-AN diagnoses despite similar BMI distributions.

To better understand the impact of weight recovery on brain features, we performed regional correlations between the residuals of each region of interest (n=82 regions) and the z-scored BMI distribution at the individual level. The thickness of 35 cortical regions and the volume of 2 subcortical regions were positively correlated with BMI in subjects with EO-AN (**Figure 2D** and Supplementary Table), showing that a z-scored BMI closest to 0 SD (=closest to a normal weight) was associated with greater CT and volume. The highest correlation was observed for the middle temporal gyrus (**Figure 2E**, r=0.37, p_adj_=0.001). In contrast, despite similar BMI distributions (Supplementary Figure 2), we found no significant correlation for the CT or volume of any region in subjects with ARFID (Supplementary Table). No correlation between SA and BMI survived FDR correction in EO-AN or ARFID.

### Structural brain similarities between EO-AN and NDDs mirror underlying genetic correlations

We extracted and ranked Cohen’s *d* distributions (**Figure 3AB**) for the brain signatures on CT of three neurodevelopmental conditions (OCD, ASD, ADHD) - and compared them with EO-AN. We did not include ARFID in the comparison as no significant results were found at the regional level when including the entire group of patients. EO-AN had the largest effect on CT (mean absolute top-decile=0.97), followed by OCD (0.25), ASD (0.21), and ADHD (0.18). Pearson correlations between EO-AN and these three conditions – as well as permutation tests, showed significant correlations between EO-AN and OCD (r_B AN-OCD_=0.59, p_perm_=0.0001). Correlations between EO-AN and ADHD, as well as ASD, were not significant (r_B AN-ADHD_=-0.14, r_B AN-ASD_ =-0.03). Lastly, we observed a concordance of 0.94 between brain-based (computed in this paper) and genetic (previously published) correlations across the three pairs of conditions (**Figure 3C**).

**Figure 3.**
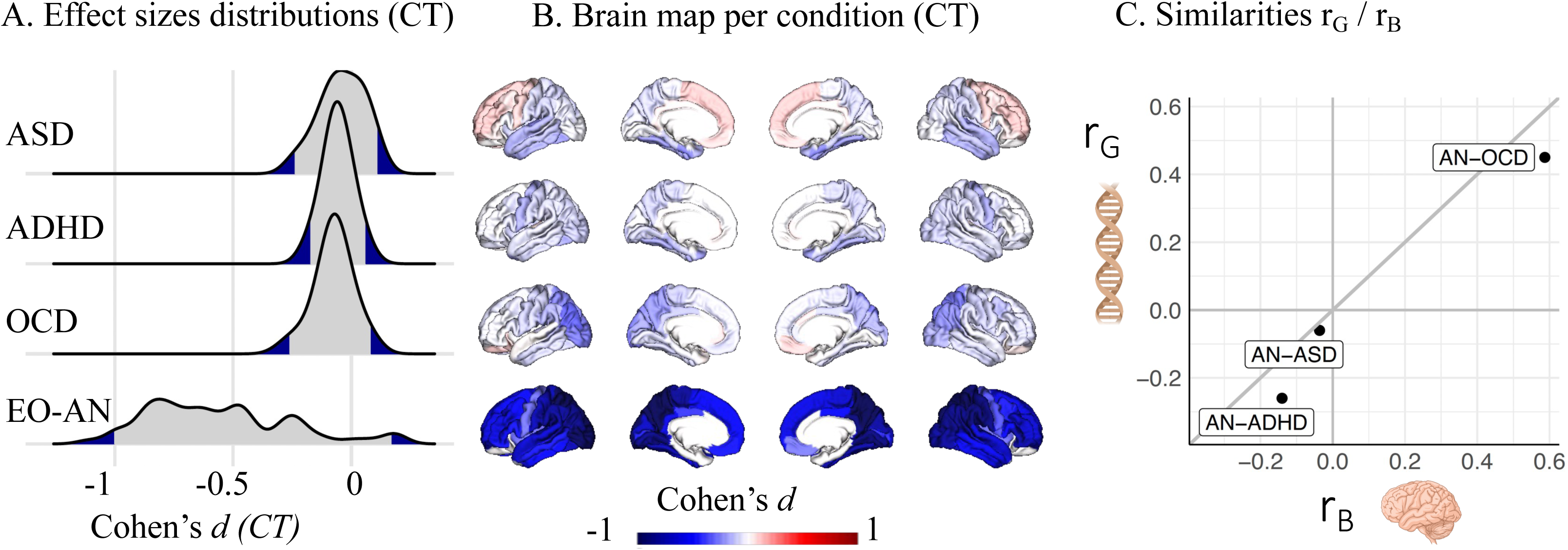
Similarities of the EO-AN brain pattern with additional psychiatric conditions. *A.* Distribution of effect sizes of three psychiatric conditions on regional CT (previously published by the ENIGMA consortium), as well as EO-AN (computed in this study). *B.* Brain maps representing patterns of abnormalities in CT reported by the ENIGMA consortium for three psychiatric conditions (see methods) and EO-AN. *C.* Relationship between SNP-based correlations (r_G_, provided by ^14,27^) and brain-based correlations (computed in this study). Abbreviations: ADHD: Attention Deficit Hyperactivity Disorder, ASD: Autism Spectrum Disorder, OCD: Obsessive-Compulsive Disorder, CT: Cortical Thickness.

## Discussion

To the best of our knowledge, we conducted the largest neuroimaging study on patients with an rEO-ED. Patients with an early onset form of AN presented a large widespread pattern of thinner cortex and smaller subcortical and total gray volumes (compared to TD), similar to results obtained on patients with a typical onset, suggesting an effect independent of the age of onset. Despite similar BMI distributions in EO-AN and ARFID, we did not identify any cortical thickness alterations in those with ARFID. However, we observed smaller intracranial and left hippocampus volumes in ARFID (compared to TD), and a large reduction of SA in the unARFID group, suggesting different mechanisms underlying both restrictive rEO-ED conditions. Half of the brain abnormalities in EO-AN were no longer observed after partial weight restoration, pointing toward a partial mediation of the BMI on the brain signal. Lastly, we identified overlapping patterns of brain differences reported in EO-AN and OCD, mirroring findings observed at the genetic and clinical levels. This multi-scale stability of the overlap points toward potential shared mechanisms underlying psychiatric disorders independent of BMI.

*Hypothesis-1* predicted that individuals with EO-AN would exhibit a widespread pattern of thinner cortex compared to TD, particularly in later-developing brain regions, such as the anterior cingulate cortex and the insula ^21^. However, our results revealed an overall effect of EO-AN on CT without a specific impact on this particular set of regions. Furthermore, EO-AN and TO-AN showed highly overlapping brain patterns compared to TD (r=0.8), indicating similar brain alterations regardless of age of onset. The only notable difference was in the effect’s amplitude: EO-AN showed larger deviations from TD than TO-AN, as well as ADHD, ASD and OCD.

Half of the cortical regions’ thickness correlated with BMI in EO-AN patients, supporting the existence of a widespread consequence of low BMI on CT, but also a specific effect of EO-AN independent of BMI. The underlying mechanisms of brain structural alteration in AN remain unclear. Partial restoration of brain thickness abnormalities following weight partial recovery has been speculated to be associated with hydration level and/or changes in oncotic pressure ^22^. To address these hypotheses, studies have investigated gravity of urine samples 1h before scanning but did not find any abnormality, excluding the likelihood of the dehydration hypothesis ^23^. Serum albumin assessment prior to scanning was also reported as normal, reducing the likelihood of an increased blood volume after weight partial recovery ^22^. Alternative hypotheses should be investigated, such as differences in metabolic costs of brain regions ^24^. BMI may not be the most appropriate measure for assessing body fat to investigate AN mechanisms. Other metrics, such as leptin levels or body fat derived from MRI data, could provide more accurate measures ^25^.

*Hypothesis-2* predicted that children and adolescents with EO-AN and ARFID would show partially overlapping brain patterns driven by low weight. Despite equivalent BMI distributions in both conditions, EO-AN and ARFID showed differential brain patterns, which suggests independent underlying mechanisms. ARFID was associated with lower intracranial and hippocampus volume than TD, but also smaller SA in the subgroup of patients with a BMI<3rd percentile. Surprisingly, we did not observe any effect on CT, and any association between BMI and any brain metrics. One hypothesis for the lack of correlation may relate to the age of onset of the condition. Children with ARFID typically experience a more gradual onset of their eating disorder compared to those with EO-AN. This is particularly the case for two out of three clinical subgroups: a lack of interest in food, and sensory sensitivity impairment. This slower progression could allow the brain to compensate for chronic low-calorie intake in children with ARFID compared to children with EO-AN ^26^.

*Hypothesis-3* predicted a significant overlap in brain alterations between EO-AN and other NDDs - mirroring previously published clinical observations and genetic results. We were able to identify such similarities at the brain level of EO-AN with OCD, ADHD (negative correlation), and ASD (no correlation), which mirror genetic risk correlations from these same disorders^14,27^. Positive correlations with OCD, as well as no association with ADHD, are consistent with clinical observations ^1,28,29^. Overall, this multi-scale overlap (at the clinical, brain, and genetic levels) points toward potential shared mechanisms underlying psychiatric disorders independent of BMI and supports the importance of large-scale transdiagnostic approaches to better understand the underlying mechanisms of mental illnesses. These results also align with the major development of multidimensional conceptualization of psychiatric conditions, such as the Research Domain Criteria (RDoC ^30^) or the general psychopathology factor (p-factor ^31^).

### Limitations

#### ARFID subtypes

Due to the limited sample size, we could not subtype patients based on the three clinical subgroups, which would be valuable in the future.

#### Cross-sectional dataset

This dataset was cross-sectional rather than longitudinal. However, each patient (diagnosed with AN or ARFID) was hospitalized with a BMI percentile <3. This indicates that those with a BMI above this threshold had partially recovered and recently gained weight. This allowed us to infer about the stages of the brain recovery process.

#### Intellectual Quotient (IQ)

No IQ testing was performed on the typically developing children.

#### Genetic and brain-based correlations

sample sizes used to build summary statistics required for these genetic-based correlations, as well as the ones used to compute brain signatures, were not always consistent, which could have introduced some biases.

## Conclusion

This large-scale neuroimaging study reveals distinct brain alterations in two rEO-EDs, highlighting unique biological mechanisms underlying each condition. Future studies will be required to partition the contribution of low BMI *vs.* rEO-ED mechanisms, as well as to identify shared mechanisms with other mental illnesses toward a dimensional approach of ED.

## Materials and Methods

### Study Samples

We analyzed data from 221 patients hospitalized in the eating disorder clinic of the psychiatric department of Robert Debré Hospital (Paris, France). Sample sizes reported in the section below are after MRI image quality control (see flowchart in Supplementary Figure S1).

#### Children with early-onset anorexia

All patients met the DSM-5 criteria for AN and were inpatients at the time of their scan. The age of first inpatient admission for nutrition rehabilitation under 13 years old was used as a criterion to categorize patients as part of the early onset subgroup of AN ^9^. Details about the inpatient program are provided in Supplementary Material. The EO-AN sample consisted of 124 children (85% females), with a mean age at first inpatient admission of 11.40 (± 1.07 SD) and a mean age at scan of 11.5 (± 1.18 SD) years (**Table 1**). We split the EO-AN patients into two subgroups based on the BMI percentile at the time of the MRI scan - to account for the disease trajectory: the acutely ill group (EO-acAN, n=60, percentile <3) and the partially weight-restored group (EO-pwrAN, n=64, percentile>=3) (**Table 1, Figure S2**). We calculated youth BMI percentile from the Centers for Disease Control and Prevention standards using the *PAutilities* R package.

**Table 1.**
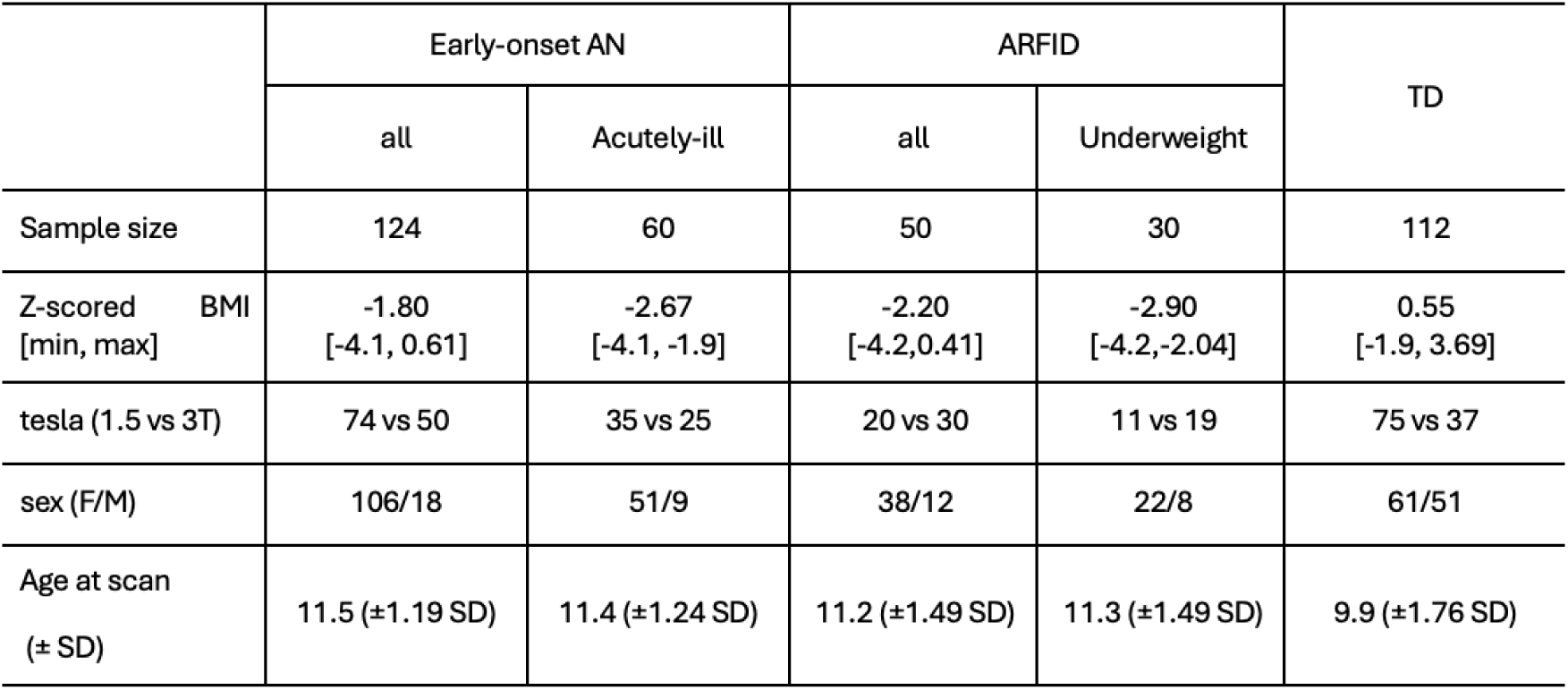
Demographic information of participants included in this study. Demographic information on inpatients with a diagnosis of early onset anorexia nervosa (EO-AN) or avoidant restrictive food intake disorder (ARFID) and children and early adolescents with a typical development (TD). The numbers reported in this Table were those that remained after quality control. For quantitative variables, data are mean [minimum, maximum] or (±standard deviation, SD). BMI: body mass index, F: female, M: male.

|  | Early-onset AN |  | ARFID |  | TD |
| --- | --- | --- | --- | --- | --- |
|  | all | Acutely-ill | all | Underweight |  |
| Sample size | 124 | 60 | 50 | 30 | 112 |
| Z-scored BMI<br>[min, max] | -1.80<br>[-4.1, 0.61] | -2.67<br>[-4.1, -1.9] | -2.20<br>[-4.2, 0.41] | -2.90<br>[-4.2, -2.04] | 0.55<br>[-1.9, 3.69] |
| tesla (1.5 vs 3T) | 74 vs 50 | 35 vs 25 | 20 vs 30 | 11 vs 19 | 75 vs 37 |
| sex (F/M) | 106/18 | 51/9 | 38/12 | 22/8 | 61/51 |
| Age at scan<br>(± SD) | 11.5 (±1.19 SD) | 11.4 (±1.24 SD) | 11.2 (±1.49 SD) | 11.3 (±1.49 SD) | 9.9 (±1.76 SD) |

#### Children with ARFID

We enrolled 50 inpatients with an ARFID diagnosis (26%, n=13 with a lack of interest in food; 58%, n=29 with fear of aversive consequences of food intake; 16%, n=8 with sensory sensitivity impairment). All ARFID patients included in this study were diagnosed based on the DSM-5 criteria for ARFID ^11^, and a BMI under the third percentile at the time of hospitalization. The mean age and sex ratio was similar to the EO-AN group (mean age at scan =11.2 [7.6, 14.7](± 1.49 SD)) years, 76% female). We subdivided the ARFID group according to each participant’s BMI percentile at the time of the scan. Participants with a BMI percentile < 3 were considered as the ‘underweight group”: *un*ARFID (n=30).

#### Children with Typically Developing

Participants (n=112) were recruited from the control groups of the Paris Autism Research International Sibpair (PARIS) and the DEVine cohorts at the Child and Adolescent Psychiatry Department, Robert Debré Hospital, Paris (France) (mean age at scan =9.9(±1.76SD) years, 54% female). TD were scanned using the same imaging scanners as the AN and ARFID.

More details about these three groups (AN, ARFID, TD) are available in Supplementary Material.

### Image Acquisition and Processing

#### Scanning Protocols

Data were collected between 2010 and 2024 on three Philips scanners at two magnetic field strengths (1.5 and 3 Tesla, Table 1). Acquisition parameters and specific sequence information are available in the Supplementary Methods section. Sensitivity analyses were performed on data collected on the 3T machine only (between 2019 and 2024) - see Supplementary results.

Brain segmentation and metrics extraction: All T1-weighted structural brain MRI scans were processed using FreeSurfer (version 7.2.0 ^32^) through the computing cluster at the Human Genetics & Cognitive Functions lab, Institut Pasteur (Paris, France). Quality control of the T1-weighted brain MR images and the segmentation of the cortical and subcortical regions was performed by the same rater (AA). Images were segmented using the Desikan-Killiany atlas ^33^ into 68 cortical and 14 subcortical regions + 2 lateral ventricles. Cortical thickness (CT) and surface area (SA) measures were extracted for the 68 cortical regions (34 per hemisphere). Volumes were calculated for the 16 subcortical regions. We also extracted global metrics from FreeSurfer: mean CT, total SA, intracranial volume (ICV), total gray volume (GV), and cerebrospinal fluid (CSF) volume.

### Statistical analyses

#### Case-Control analysis

Group difference analyses (EO-AN>TD, ARFID>TD, EO-acAN>unARFID, EO-acAN>TD, unARFID>TD) were performed to assess the impact of rEO-ED diagnosis on brain metrics. We used *ComBat* batch adjustment method ^34^, which has been extensively used in neuroimaging analyses to reduce site-related heterogeneity and increase statistical power for multisite analysis. We treated each sequence-machine pairing as an individual site. Batch-adjusted analyses were conducted using *lm* function, to investigate the effect of the rEO-ED diagnostic on brain metrics, using sex and age at scan as covariates. We applied the False Discovery Rate (FDR) procedure ^35^ to correct for multiple testing. Analyses of the subcortical volumes were adjusted for intracranial volume (ICV). CT analyses reported in the main text were not adjusted for global metrics. CT results adjusted for mean thickness were reported in Supplementary Results.

#### Brain profiles correlation with TO-AN

Analyses comparing EO- and TO-AN were performed using previously published TO-AN data from the ENIGMA Eating Disorders working group ^10^. Additional information on this sample is provided in Supplementary Material.

#### Concordance analyses between brain-based and genetic-based correlations

##### Brain structural patterns for three neurodevelopmental conditions

We extracted 84 Cohen’s *d* values (for 68 cortical and 16 subcortical regions) for each of the three following psychiatric conditions (published data from the ENIGMA consortium): attention deficit hyperactivity disorder (ADHD, n=1,513 cases *vs.* 1,395 controls) ^36,37^, ASD (1,571 cases *vs.* 1,651 controls) ^38^ and OCD (n=407 cases *vs.* 324 controls) ^39,40^. Results from each of these studies were derived from the largest, multisite cohorts ever analyzed at the time - and were analyzed using the same, publicly available ENIGMA processing pipelines.

##### Brain-based correlations across disorders

Recent analytic methods allow resemblance among brain maps to be tested formally, allowing affinities among different conditions to be evaluated ^41^. We computed Pearson correlations between beta value vectors for each pair of conditions. To avoid inflation of spatial correlations, we performed spin permutation tests (5,000 null correlations per pair of brain maps) proposed by ^41^ and implemented in the ENIGMA toolbox ^42^.

##### Transdiagnostic genetic risk correlations

We extracted genetic correlations across the same three pairs of conditions computed by the Psychiatric Genomics Consortium (PGC) ^14,27^ using the largest genome-wide association studies per condition (GWAS).

### Ethics

The study was carried out in accordance with the principles of the Declaration of Helsinki and followed the Good Clinical Practice (ICH GCP) standards.

Data from patients with ED (AN and ARFID) were studied following the French regulation (MR-004) and approved by the local Ethics Committee (ref:CEER 2022-610ter).

Data from the TD group were used under authorizations of the PARIS (ref: Inserm C07-33 - CEER 2008-A00019-46) and DEVine cohorts (ref: CCP 3772-RM, Protocol CEA 100 054).

All participants and their guardians provided written informed consent.

## Supporting information

Supplementary Material, Method, and Results

## Data Availability

All derived imaging data produced in the present study are available upon reasonable request to the authors

## Funding

The authors would like to thank all participants and their families who participated in the study, as well as the Clinical Investigation Center, Robert Debré Hospital, Paris, France, for managing the study. This study was supported by funding from the Fondation de France (2015-00059547) and the Institut Pasteur (Université de Paris, CNRS UMR 3571, “Contrat interface hospitalier”). CAM, PMT, SE, and RD acknowledge fundings from the National Institute of Mental Health (U01MH136221).

This work was supported by the German Research Foundation (SFB 940 TP C03, EH 367/5-1, EH 367/7-1).

## Financial Disclosure

CAM, AA, RB, AM, CS, PB, NT, LT, DG, MA, MEB, SE, TB, and RD reported no biomedical financial interests or potential conflicts of interest.

## Author contributions

CAM and AA designed the overall study, processed the imaging data, and drafted the manuscript.

AA collected and performed quality control of the imaging data, and drafted the manuscript.

RD and TB designed the overall study, contributed to the interpretation of the data, and reviewed the manuscript.

RB, AM, LT, NT, and PB processed the imaging data.

DG, MA, ME and CS collected the imaging data and contributed to the interpretation of the data.

CRKC, SE, and PMT contributed to the interpretation of the data and reviewed the manuscript.

## Notes

### Competing Interest Statement

The authors have declared no competing interest.

### Author Declarations

The study followed the principles of the Declaration of Helsinki and the Good Clinical Practice (ICH GCP) standards. Data from patients with eating disorders were studied following the French regulation (MR-004) and approved by the local Ethics Committee (CEER - 'Comite d'Evaluation de l'Ethique des Projets de Recherche' of the Robert Debre hospital: 2022-610ter). Data from the control group were used under authorizations of the PARIS (ref: Inserm C07-33 - CEER of Robert Debre hospital 2008-A00019-46) and DEVine cohorts (ref: CCP 3772-RM, Protocol 'Commissariat aux Energies Atomique et Alternative' number: 100 054). All participants and their guardians provided written informed consent.

