## Supplementary Material, Method, and Results for "Neuroimaging Insights into Brain Mechanisms of Early-onset Restrictive Eating Disorders"

Supplementary Material, Method, and Results 1
- Supplementary Material and Method 2
  - Children and adolescents with early-onset anorexia 2
  - Children and adolescents with ARFID 2
  - Children with Typically Developing 3
  - Flowchart of the participants 3
  - BMI distribution per group 4
  - Inpatient treatment and inclusion criteria 4
  - 3D T1-weighted sequences 5
  - Brain profiles correlation with TO-AN 5
- Supplementary Results 6
  - CT results adjusted for global effects. 6
  - Differences between TO-AN > TD and EO-AN > TD 7
  - Effect of unARFID on Global Brain Measures 7
  - Effect of acutely ill vs partially weight-restored patients with EO-AN on brain structure 8
  - Results on data from the 3T machine only 8
  - CT results excluding a subgroup of controls 8
  - Analyses excluding males 8
  - ARFID Subtypes 9
- Supplementary Tables 9
  - Aim 1. Case-control (EO-AN>TD) analyses - adjusted and unadjusted for meanCT 9
  - Aim 2. Case-control (ARFID>TD) analyses 9
  - Aim 2. Regional correlations between the residuals of each brain structural metric and the z-scored BMI distribution at the individual level 9
- References 9

### Supplementary Material and Method

#### *Children and adolescents with early-onset anorexia*

18.6% of the EO-AN patients were being medicated with either fluoxetine (Selective Serotonin Reuptake Inhibitor (SSRI), n=14, 2 EO-acAN / 12 EO-pwrAN), olanzapine/aripiprazole (neuroleptic, n=7, 3 EO-acAN / 4 EO-pwrAN), or both (n=2, 1 EO-acAN / 1 EO-pwrAN). We collected puberty stages using the Tanner index (assessed by a pediatric endocrinologist) for 76% of this group. One girl included in this study had reached complete puberty during hospitalization (Tanner index=5). The Wechsler Intelligence Scale for Children (WISC; fourth edition) was used to calculate the Perceptual Reasoning Index (PRI) after weight restoration [3]. Patients with EO-AN had a mean PRI=114[77,148](38NA). Given that our research sample spanned a long period of time (between 2010 and 2024), patients were diagnosed according to either the DSM IV-TR or the DSM-5 criteria for AN [1,2], including a BMI under the third percentile at the hospitalization time. To ensure consistency, we performed a diagnosis conversion from DSM IV-TR to DSM-5 based on clinical records. No patient had purging behavior or binge eating (restrictive subtype).

#### *Children and adolescents with ARFID*

To meet DSM-5 diagnosis criteria for ARFID, patients must be reported with criterion A: an eating or feeding disturbance (e.g., lack of interest in food; fear of aversive consequences of food intake; avoidance based on sensory sensitivity characteristics) associated with at least one of four consequences of restrictive eating: weight loss/growth impairment (A1); nutritional deficiency (A2); dependence on enteral feeding or oral supplements (A3); and/or psychosocial functioning interference (A4). Eating disturbance should not be better explained by lack of food availability (criterion B), associated with body image concerns (criterion C), or attributable to another medical condition or mental disorder (criterion D) [1]. Psychiatrists (AA, CS) also used a semi-structured interview with the child and the parents (or caregivers) based on the DSM-5 criteria to assess the ARFID diagnosis at the hospitalization admission to improve the robustness of the diagnosis.

24% of them were medicated with either fluoxetine (n=10, five with a BMI percentile < 3, five with a BMI percentile >3), olanzapine/aripiprazole (neuroleptic, n=2, only in patients with a BMI percentile < 3). The Wechsler Intelligence Scale for Children (WISC; fourth edition) was used to calculate the Perceptual Reasoning Index (PRI) after weight restoration [3]. Patients with ARFID had a mean PRI of 108 [86, 132](26NA).

*Children with Typically Developing*

Participants were recruited from the Paris Autism Research International Sibpair (PARIS) consortium cohort at the Child and Adolescent Psychiatry Department, Robert Debré Hospital, Paris (France), including ASD patients, siblings, and TD children. We only included TD children (n=92) and ASD siblings (n=20). Sensitivity analyses were performed by excluding the subgroup of siblings and did not reveal any differences (see Supplementary Results below). We harmonized TD and patients based on age to avoid results being confounded by any developmentally associated effects (mean age at scan TD = 9.9 TD [6.2, 12.9]). Height values were unfortunately not collected for a subset of TD participants, but weight was systematically collected.

*Flowchart of the participants*

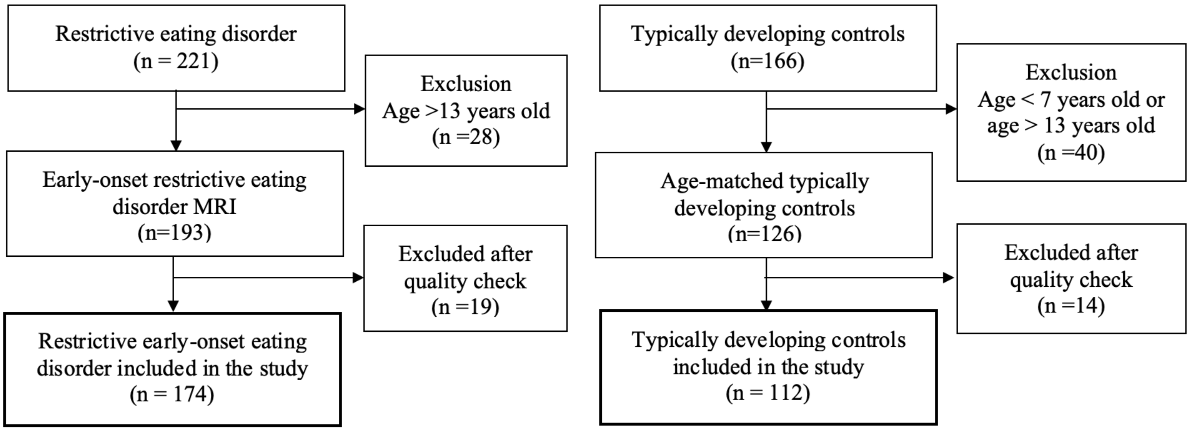


***Figure S1.*** *Flowchart of the participants.*

*BMI distribution per group*

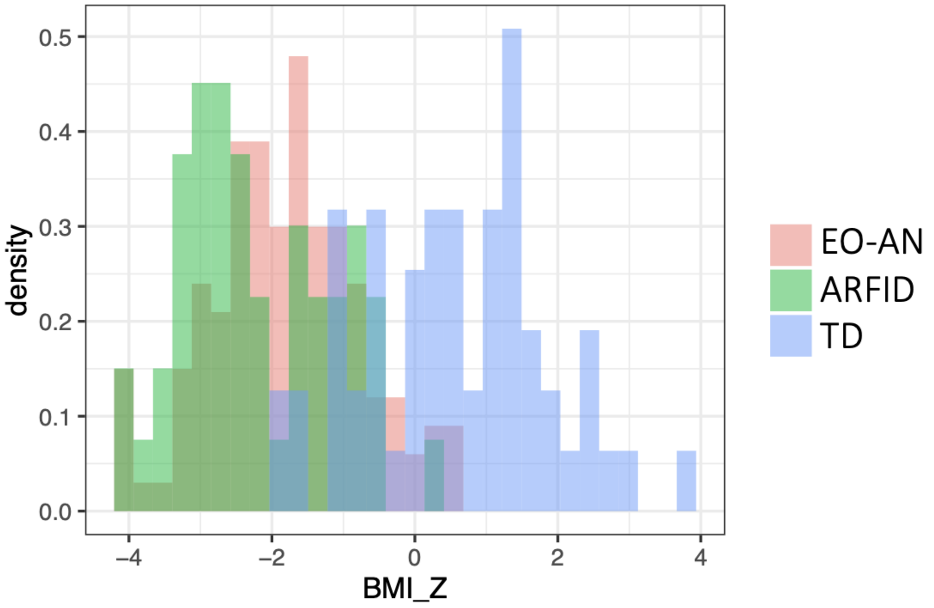


***Figure S2.*** *BMI percentile distributions of EO-AN patients (pink), ARFID patients (green) and typically developing individuals (TD, blue).*

*Inpatient treatment and inclusion criteria*

During inpatient treatment, the main nutritional goal for patients was to achieve a "healthy" weight that allows normal growth and pubertal development. The impact of psychiatric and somatic comorbidities was systematically assessed and taken into account during the treatment process.

An individualized weight restoration target was computed for each child based on age, height, stage of puberty, and growth charts (following recommendations from the Canadian Pediatric Society)[4]. Weight restoration was achieved using oral nutritional supplements or nasogastric tube feeding when nutritional needs were not met. Food intakes were gradually increased (200 kcal increments) to reach a standard weight gain target between 500g and 1kg per week until the individualized target BMI percentile was reached. Regular monitoring of electrolytes and clinical status was performed and appropriate supplementation was provided (mainly phosphorus, vitamin D, and calcium supplementations). The prescription of medication, when indicated (neuroleptics or antidepressants) was introduced after sufficient weight gain (mainly after reaching the 3rd BMI percentile).

Robert Debré Hospital (Paris, France) is a reference center for the treatment of early-onset eating disorders, and patients also undergo systematic paraclinical evaluation to search for differential medical diagnoses (such as brain tumors). A systematic MRI scan was prescribed for all patients upon their arrival at the hospital as part of a clinical routine and was performed in the days or weeks following admission based on the capabilities of the radiology department. Scans were performed during hospitalization for anorexia nervosa (AN). Patients and relevant family members provided written informed consent to participate in the study, including their brain MRI and clinical characteristics. Exclusion criteria: Participants did not receive brain MRI if 1) the patient had already received an MRI upon a visit to another hospital and 2) the presence of dental brackets/braces.

##### *3D T1-weighted sequences*

Two types of T1-weighted sequences were used:

- Fast Field Echo (FFE) sequence on a 1.5 T Ingenia Philips machine: TR = 25 ms, TE = 5.6 ms, FOV = 240x240 mm, a 1-mm isotropic voxel resolution, and 170 slices. This sequence was adapted for the 3 T Ingenia machine.
- 3D Turbo Field Echo (TFE) sequences: These were implemented on all machines with shorter TR and TE values (TR = [7-8] ms and TE = [3-4] ms), which allowed a wider FOV = 260x260 mm, 175 slices while keeping a 1mm isotropic resolution. They were also implemented on the 3T machine, with a finer voxel resolution of 0.5x0.5x0.75 mm (using Sense and Compressed Sensing (CS) to fasten the acquisition).
  An updated TFE sequence version has been recently implemented with the following parameters: TR = 6.8 ms, TE = 3 ms, FOV = 256x256 mm, 200 slices, and 0.9 mm isotropic voxel resolution.

##### *Brain profiles correlation with TO-AN*

Analyses comparing EO- and TO-AN were performed using previously published data from the ENIGMA Eating Disorders working group [5] on n=685 females with AN (mean age at scan = 21 [15-27] years, and <10th age-adjusted BMI percentile) and 963 TD individuals (**Figure 1.c**). Diagnoses were assessed based on DSM IV-TR, DSM-5, or ICD-10 criteria for AN. Case-control analyses were performed using linear regression. Models were adjusted for scanning sites using ComBat batch adjustment method. We extracted vectors of Cohen's *d* values from their case-control analyses (TO-AN>TD) for CT and subcortical volumes.

#### Supplementary Results

*CT results adjusted for global effects.*

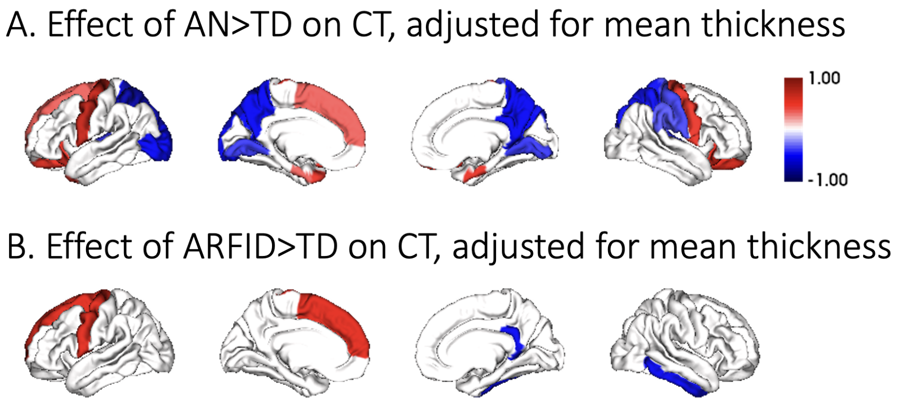


***Figure S3.*** *Effect (Cohen's d) of EO-AN versus TD (A) and ARFID versus TD (B) on CT, adjusted for mean thickness.*

*Abbreviations: ARFID: Avoidance/Restrictive Food Intake Disorder, CT: Cortical Thickness, AN: Anorexia Nervosa, TD: Typical Development, CT: cortical thickness.*

*Differences between TO-AN > TD and EO-AN > TD*

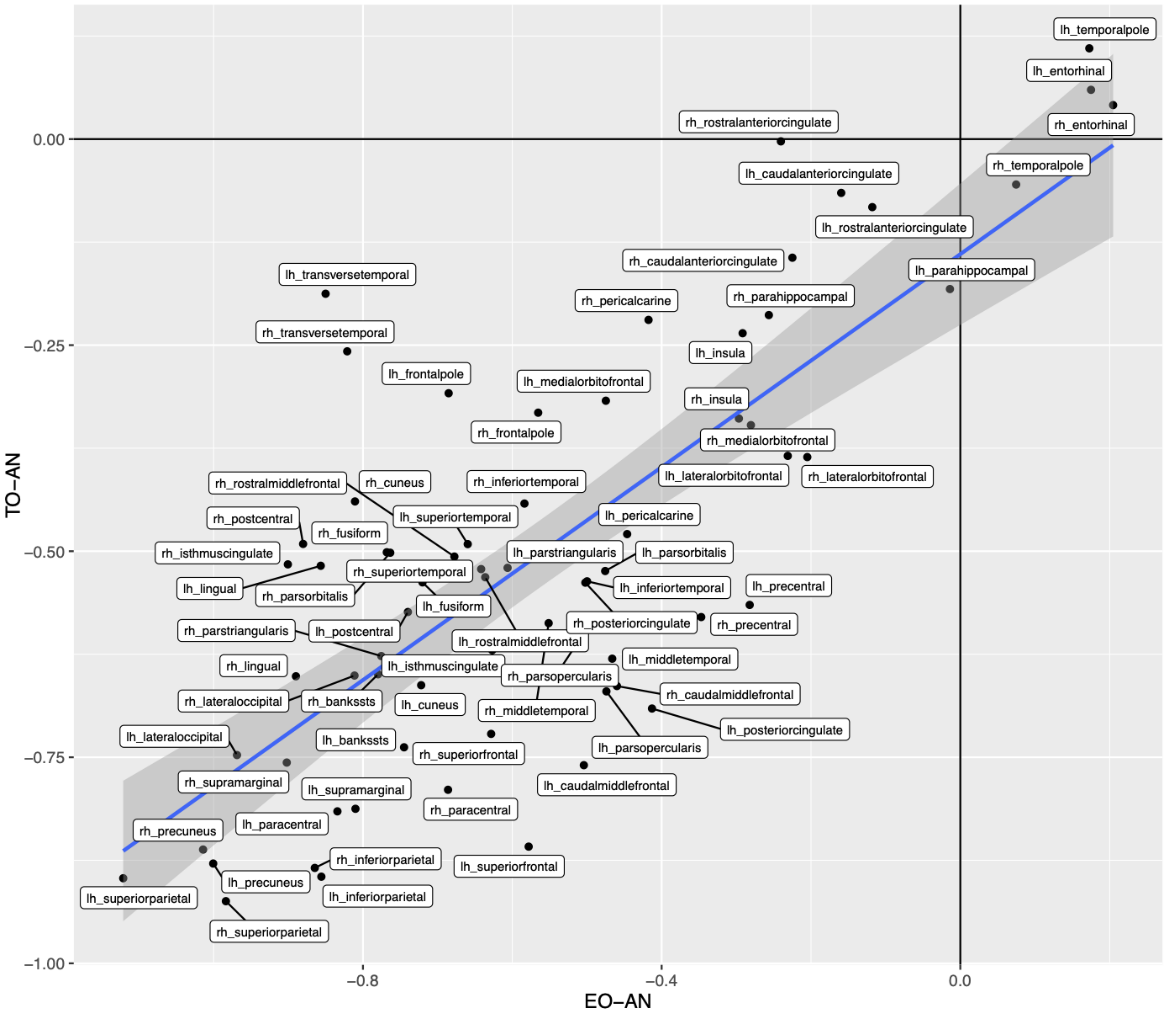


***Figure S4.*** *Effect of EO-AN (x-axis) vs. TO-AN on cortical thickness of 68 regions. EO-AN: early-onset anorexia nervosa, TO-AN: typical-onset anorexia nervosa.*

*Effect of unARFID on Global Brain Measures*

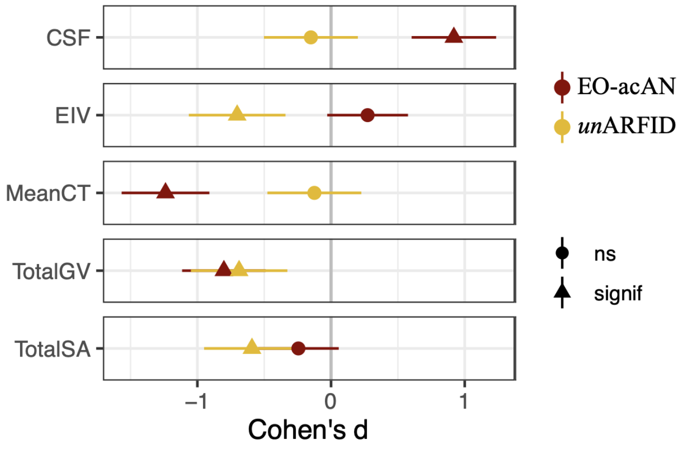

***Figure S5.*** *Effect of EO-acAN (dark red dots) and unARFID (yellow dots) on five global brain metrics (ICV: intracranial volume, Total SA: total surface area, Mean CT: mean cortical thickness, CSF: cerebrospinal fluid, Total GV: total gray matter volume). Triangles represent significant effect sizes.*

##### *Effect of acutely ill vs partially weight-restored patients with EO-AN on brain structure*

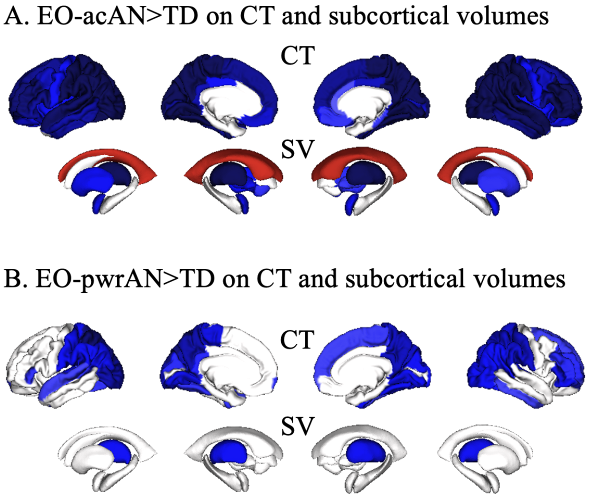


***Figure S6.*** *Effect of acute vs. partially weight-restored patients with EO-AN on brain structure Legend: Brain maps showing Cohen's d values for each of the 68 cortical regions (thickness) and the 16 subcortical regions and ventricles (volumes) for A) Acutely ill EO-AN versus TD effects on CT and subcortical volumes and B) Partially weight restored EO-AN versus TD effects on CT and subcortical volumes. Abbreviations:CT: Cortical Thickness, EO-AN: Early-Onset Anorexia Nervosa, TD: Typical Development.*

##### *Results on data from the 3T machine only*

We performed a group comparison of AN cases and TD scanned on the 3T MRI machine only (50 cases, 37 TD). Results were similar to the ones obtained using the entire sample (r=0.91, p<2.2e-16).

##### *CT results excluding a subgroup of controls*

The impact of EO-AN (n=124) versus TD (n=92), after removing ASD siblings (n=20) yielded similar results for CT (r=0.99, p<2.2e-16) and subcortical volumes (r=0.98, p=1.9e-14) as those obtained when the entire control group was included (n=112).

##### *Analyses excluding males*

Findings obtained when conducting case-control analyses on females only (n=106 AN vs. n=61 TD) closely mirrored those obtained when analyzing males and females together (r=0.97).

##### *ARFID Subtypes*

The ARFID subgroup of 'fear of aversive somatic consequences of food intake' is reported to have an increased likelihood of weight loss and a shorter length of illness [6]. If a part of the reduction of CT observed in AN is due to rapid weight loss, we should observe a similar abnormality pattern in this ARFID subgroup. We performed a case-control analysis with this ARFID behavioral subtype ('Fear of aversive consequences,' n=29). Before adjusting for multiple comparison corrections, we observed lower thickness in the right fusiform gyrus (Cohen's $d$=-0.48, p=0.03). No region survived FDR correction.
